# Causal Relationship Between Metformin and the Abundance of Ten Intestinal Genera-Level Bacterial Populations: A Mendelian Randomization Analysis

**DOI:** 10.1101/2025.07.10.25331293

**Authors:** Xiaoyong Wang, Ningning Ma, Qi Chen, Yujian Wu

## Abstract

**Objective:** This study aims to investigate the causal effects of metformin on the abundance of 10 gut bacteria genera using Mendelian randomization(MR) analysis. The genera examined include *Parabacteroides, Bacteroides, Alistipes, Adlercreutzia, Anaerofilum, Oxalobacter, Ruminococcaceae UCG003, Ruminococcus1, Butyrivibrio*, and *Eubacterium oxidoreducens* group.

**Methods:** Data from public databases were utilized, with metformin as the exposure variable and the abundance of the 10 gut bacteria genera as outcome variables. An inverse variance-weighted method was employed for MR analysis to assess the causal effects of metformin on gut bacterial abundance using genetic instrumental variables.

**Results:** The analysis revealed significant causal associations between metformin and the abundance of multiple gut bacteria genera. Specifically, metformin was positively correlated with *Parabacteroides, Bacteroides, Alistipes, Adlercreutzia, Anaerofilum, Oxalobacter, Ruminococcaceae UCG003*, and *Ruminococcus1*. Conversely, metformin was negatively correlated with *Butyrivibrio* and *Eubacterium oxidoreducens* group. These findings suggest that metformin may exert its pharmacological effects by modulating the abundance of gut bacteria.

**Conclusions:** There are significant causal associations between metformin and the abundance of multiple gut bacteria genera, indicating that the gut microbiota may be an important target for the pharmacological effects of metformin. This discovery provides new insights for further research into the mechanisms of metformin and the relationship between the gut microbiota and diseases.

## Background

Since its introduction into clinical practice in 1957, metformin has established itself as the preferred first-line treatment for type 2 diabetes worldwide, owing to its significant glucose-lowering effects, favorable safety profile, and low cost [1]. In addition to its classic glucose-lowering mechanisms, such as improving insulin resistance and inhibiting hepatic glucose output, metformin also exhibits multiple non-glucose-lowering benefits, including cardiovascular protection, anti-tumor effects, antioxidant and anti-inflammatory properties, improvement of polycystic ovary syndrome, weight reduction, and a decreased risk of dementia [2, 3]. However, the complex processes through which metformin exerts its pharmacological effects in vivo have not been fully elucidated. This lack of clarity regarding its mechanisms limits our ability to accurately predict and optimize the therapeutic application of metformin.

The human gut microbiome comprises at least 1,800 genera and 15,000–36,000 bacterial species, with the majority belonging to the phyla Firmicutes (65.7%), Bacteroidetes (16.3%), Proteobacteria (8.8%), and Actinobacteria (4.7%) [4]. The gastrointestinal microbiota is typically categorized into transient (allochthonous) and resident (autochthonous) communities. The resident microbiota consists of permanent microorganisms inhabiting specific niches within the gastrointestinal tract, primarily composed of two anaerobic phyla, Firmicutes and Bacteroidetes, which are involved in nutrient absorption in the intestinal mucosa. As a vast and complex ecosystem within the human body, the gut microbiota profoundly influences metabolic health by participating in various physiological processes, including host energy metabolism, short-chain fatty acid production, and inflammation regulation [5]. Increasing evidence indicates that dysbiosis of the gut microbiota is closely associated with the development and progression of numerous metabolic diseases, such as obesity, diabetes, and inflammatory bowel disease [6]. Consequently, the gut microbiota has emerged as a critical target for metabolic disease research, and modulating its composition and function holds promise for novel preventive and therapeutic strategies for these conditions.

In recent years, epidemiological studies have revealed significant differences in gut microbiota composition between metformin users and non-users, including alterations in the ratio of Bacteroidetes to Firmicutes and fluctuations in the abundance of specific butyrate-producing bacterial genera [7]. These findings suggest that metformin may exert its pharmacological effects by modulating the gut microbiota. However, observational studies struggle to distinguish causality from confounding bias, as factors such as diet and genetic background may simultaneously influence both metformin use and gut microbiota composition, thereby obscuring direct causal relationships between metformin and the gut microbiota.

Mendelian Randomization (MR) studies utilize genetic variants associated with exposure factors (e.g., drug use) as instrumental variables (IVs) to effectively circumvent issues of reverse causation and confounding inherent in traditional observational studies [8], providing an ideal framework for exploring causal drug-microbiota effects. Currently, research on metformin and the gut microbiota is still in its infancy, and systematic analyses of causal relationships at the genus level remain lacking. By systematically evaluating the causal effects of metformin on the abundance of specific bacterial genera, we can gain a more precise understanding of how metformin influences metabolic health through gut microbiota modulation. This not only aids in elucidating the mechanisms of metformin action and optimizing its clinical application beyond diabetes but also uncovers potential roles of the gut microbiota, offering novel insights and approaches for treating related diseases.

## 1. Methods and Data

### 1.1 Study Design

This study employed MR for secondary data analysis using existing databases. MR is a method that identifies causal relationships between exposure phenotypes and outcomes by utilizing genetic variants associated with the exposure as IVs. In this study, the exposure factor was metformin (Met) use, and the outcome variable was the abundance of gut bacterial genera. The study design was based on the following three core assumptions:

1. Relevance Assumption: Genetic variants (Single Nucleotide Polymorphisms, SNPs) must exhibit a strong association with the exposure factor to ensure that the selected SNPs serve as valid proxies for the exposure. To identify SNPs strongly associated with the exposure factor, this study adopted a significance threshold of p < 5 ×10^−8^. Subsequent analyses further validated the robustness of the results through sensitivity analyses and heterogeneity tests. The F-statistic was calculated to estimate sample overlap effects and weak instrument bias, using the formula: F = [formula not fully provided in the original text, typically 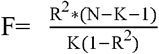, where 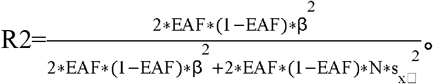 Here, N represents the sample size of the exposure factor, K is the number of IVs included, R^2^ denotes the proportion of variance in the exposure factor explained by the IVs, EAF is the effect allele frequency, β is the effect size, and SE(β) is the standard error of β. An F-statistic > 10 was required [9].
2. Independence Assumption: IVs must be independent of any known or unknown confounding factors to avoid interference from confounders and ensure accurate causal inference. During the data screening phase, we adhered to the independence principle of MR analysis by ensuring that exposure and outcome data were derived from independent samples to prevent sample overlap bias. In the data preprocessing phase, palindromic SNPs with intermediate allele frequencies were removed. Additionally, we set a linkage disequilibrium (LD) threshold of r^2^ = 0.001 and a physical distance threshold of kb = 10 Mb to ensure the independence of the IVs.
3. Exclusion Restriction Assumption: SNPs should influence the outcome solely through the exposure factor and not through other direct causal pathways. This study comprehensively validated the validity of the exclusion restriction assumption through horizontal pleiotropy tests, heterogeneity assessments, and sensitivity analyses, thereby ensuring the accuracy and robustness of causal inferences regarding the relationship between Met use and gut microbiota abundance.

These steps collectively ensured that the IVs were strongly associated with the exposure factor and independent of each other, thereby enhancing the accuracy and reliability of the MR study.

### 1.2 Statistical Analysis Methods

This two-sample MR analysis was conducted using R software (version 4.4.2) and the TwoSampleMR package (version 0.6.8). In this study, the inverse variance-weighted fixed-effects model (IVW-FE) was selected as the primary MR analysis method. Additionally, several other methods were employed to estimate (support the estimation of) causal effects, including the weighted median (WMed), MR-Egger regression (MER), simple mode (SM), weighted mode (WM), and inverse variance-weighted random-effects model (IVW-RE). The WMed, MER, SM, WM, and IVW-RE methods were used to support the evaluation of MR effect sizes. Consistent effect size directions (β values) across different methods with those obtained from IVW indicated robust results.

The mr() function from the TwoSampleMR package was used for MR analysis. To ensure statistical power, only two-sample datasets containing more than 10 eligible SNPs were included. For each eligible two-sample dataset, the log odds ratio (LogOR) and its 95% confidence interval (CI) were calculated to assess the causal association between the exposure factor and the outcome. Simultaneously, the statistical test module of the mr() function was used to compute the P-value corresponding to the LogOR to evaluate the significance of the causal association. A P-value < 0.05 was considered statistically significant, indicating a potential causal association between the exposure factor and the outcome. The magnitude of the LogOR and its 95% CI reflected the strength of the association, with a positive LogOR indicating a positive correlation and a negative LogOR indicating a negative correlation between the exposure factor and the outcome.

The Cochran Q test was used to assess heterogeneity, with a Q-statistic P-value > 0.05 indicating no significant heterogeneity [10]. Horizontal pleiotropy was tested and calibrated using the intercept and P-value from MR-Egger regression, with a P-value > 0.05 suggesting no significant horizontal pleiotropy. Two-sample datasets exhibiting significant heterogeneity or horizontal pleiotropy were excluded to reduce bias, enhance result robustness, and satisfy the fundamental assumptions of MR. Additionally, a leave-one-out (LOO) sensitivity analysis was performed to evaluate result stability by sequentially excluding each IV.

### 1.3 Data Sources

The data for this study were obtained from the IEU OpenGWAS project (https://gwas.mrcieu.ac.uk/), which provides extensive GWAS summary data for MR analysis. Metformin use was selected as the exposure factor (GWAS ID: ukb-b-14609). Outcome factors were chosen based on the “trait” item in the GWAS database, searching for GWAS datasets related to gut microbiota abundance. Based on the aforementioned study design and statistical methods, MR analysis was performed using the exposure factor ID and datasets related to gut microbiota abundance. Ultimately, 10 gut bacterial genera at the genus level were selected: *Parabacteroides, Bacteroides, Alistipes, Adlercreutzia, Anaerofilum, Oxalobacter, Ruminococcaceae UCG003, Ruminococcus 1, Butyrivibrio*, and *Eubacterium oxidoreducens group* (a genus-level related group). Details of the exposure and outcome datasets are provided in Table 1. As the data were sourced from publicly available GWAS databases, no additional ethical approval was required.

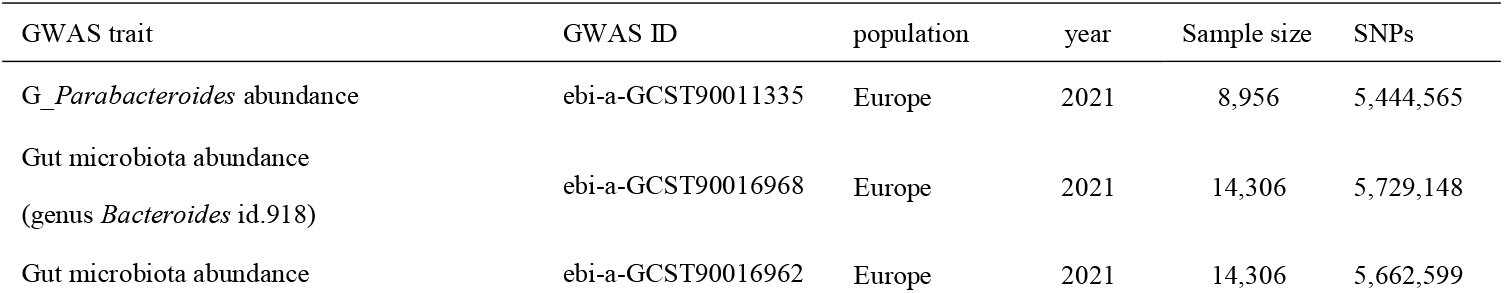
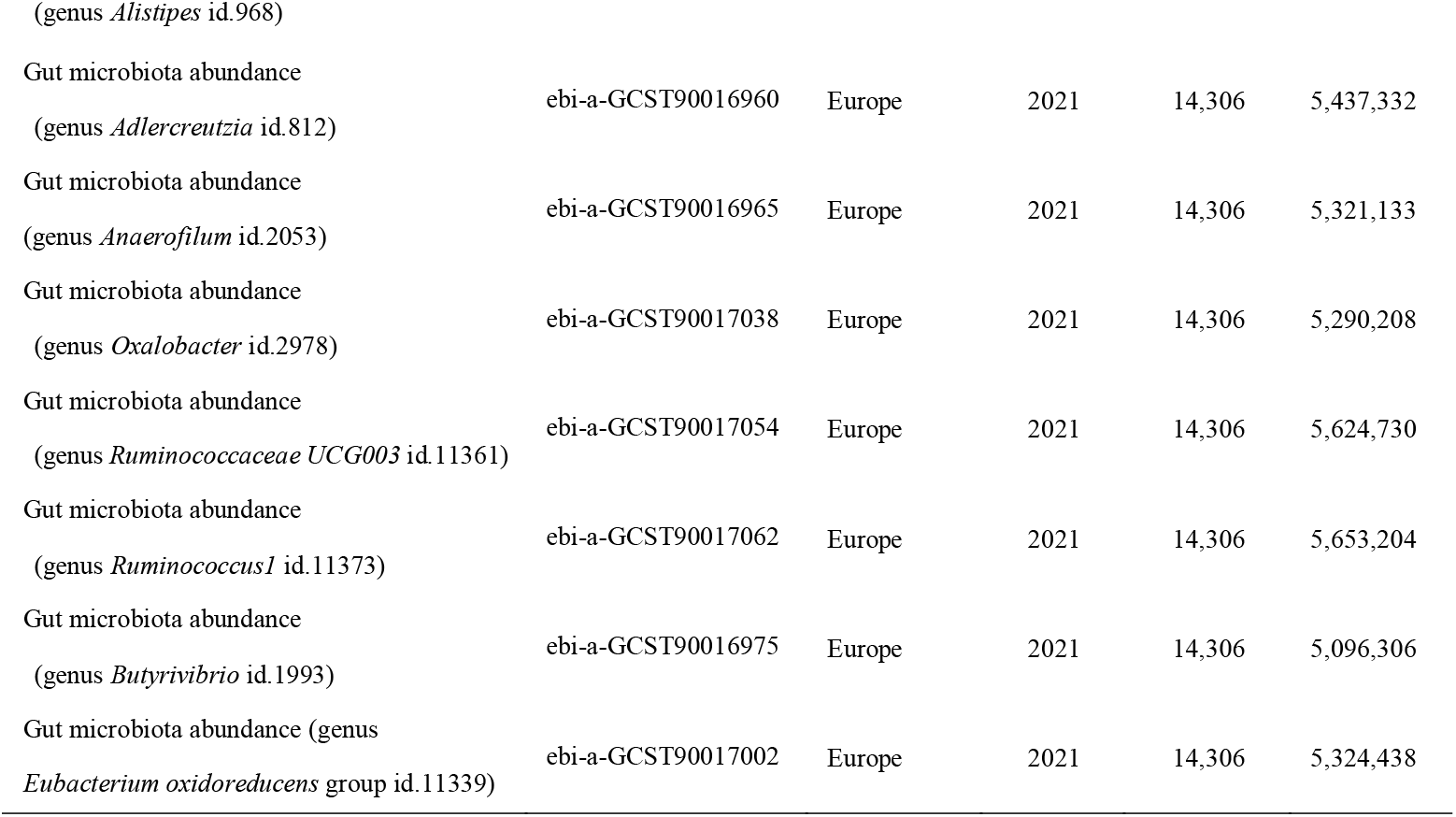

## 2 Results

### 2.1 Met showed significant positive causal associations with the abundance of the following eight intestinal microbiota

When analyzing the association between SNPs and Met, the calculated F-values ranged from 30.25 to 577.91. Based on the IVW-FE analysis method, significant positive causal associations were found between Met and eight intestinal microbiota: *Parabacteroides* (logOR = 3.812, 95% CI: 1.828–5.796, P < 0.001), *Bacteroides* (logOR = 1.473, 95% CI: 0.079–2.866, P = 0.038), *Alistipes* (logOR = 2.131, 95% CI: 0.598–3.663, P = 0.0064), *Adlercreutzia* (logOR = 2.029, 95% CI: 0.135–3.924, P = 0.036), *Anaerofilum* (logOR = 2.766, 95% CI: 0.325–5.206, P = 0.026), *Oxalobacter* (logOR = 2.879, 95% CI: 0.225–5.533, P = 0.034), *Ruminococcaceae UCG003* (logOR = 1.712, 95% CI: 0.085–3.339, P = 0.039), and *Ruminococcus 1* (logOR = 1.828, 95% CI: 0.465–3.191, P = 0.009). The Q-statistic indicated no significant evidence of heterogeneity (P > 0.05), and no pleiotropy was detected in the MR-Egger regression analysis (P > 0.05) (see Figure 1). The MR scatter plot suggested consistent directions. During the LOO sensitivity test, after removing some individual SNPs, the confidence intervals for *Bacteroides, Adlercreutzia, Anaerofilum*, and *Ruminococcaceae UCG003* included zero, indicating less stable results. The β-value directions from five analysis methods (WMed, MER, SM, WM, IVW-RE) were consistent with those from IVW-FE, indicating robust results. The funnel plot analysis showed no significant asymmetry, suggesting no publication bias.

**Figure 1.**
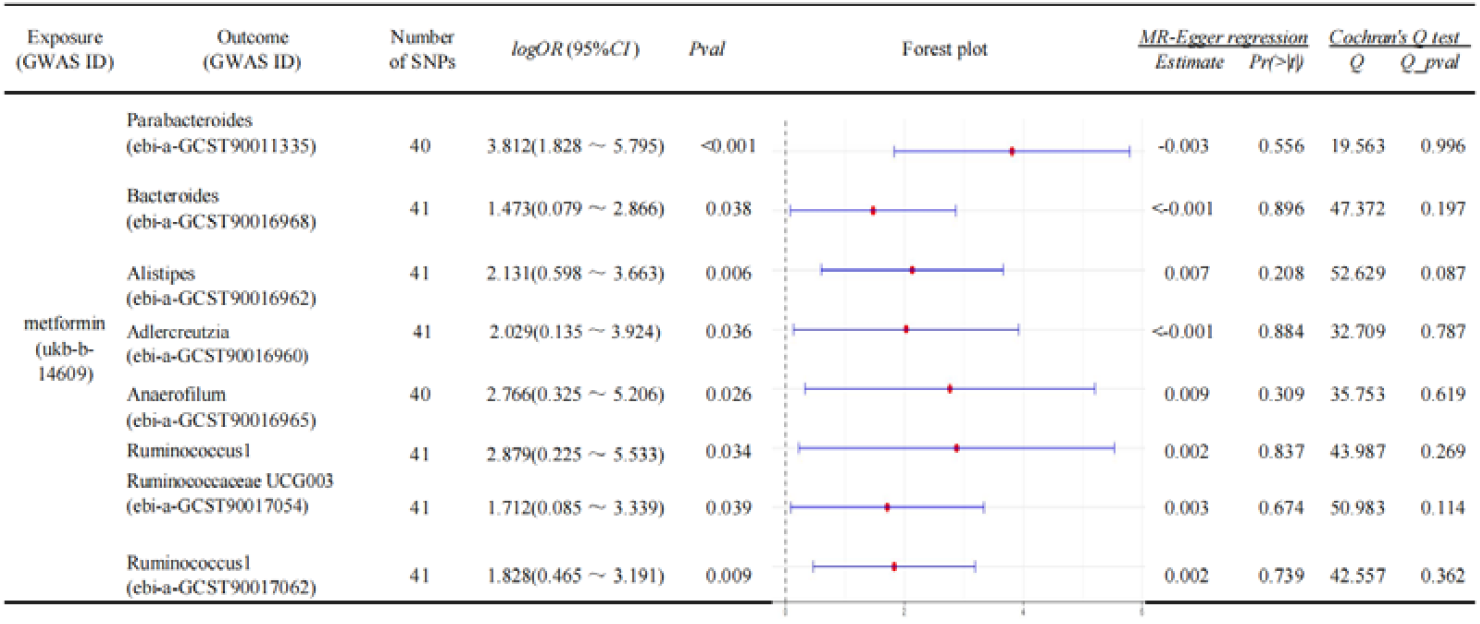
Based on the IVW-FE analysis method, the analysis of the associations between Met and eight intestinal microbiota, including *Parabacteroides, Bacteroides, Alistipes, Adlercreutzia, Anaerofilum, Oxalobacter, Ruminococcaceae UCG003*, and *Ruminococcus 1*, included the number of SNPs, forest plots of associations, logOR, 95% CI, P-values (pval), intercepts from MR-Egger regression and corresponding horizontal pleiotropy P-values (Pr(>|t|)), and Q-values and corresponding P-values (Q_pval) from Cochran’s Q heterogeneity test.

### 2.2 Met showed significant positive causal associations with the abundance of the following two intestinal microbiota

When analyzing the association between SNPs and Met, the calculated F-values ranged from 30.25 to 577.91. Based on the IVW-FE analysis method, significant negative causal associations were found between Met and two intestinal microbiota: *Butyrivibrio* (logOR = -3.513, 95% CI: -6.447 to -0.580, P = 0.019) and *Eubacterium oxidoreducens* groups (logOR = -3.562, 95% CI: -5.936 to -1.187, P = 0.0033) (P < 0.05). The Q-statistic indicated no significant evidence of heterogeneity (P > 0.05), and no pleiotropy was detected in the MR-Egger regression analysis (P > 0.05) (see Figure 2). The MR scatter plot suggested consistent directions (see Figure 3). The LOO sensitivity test indicated robust results. The β-value directions from five analysis methods (WMed, MER, SM, WM, IVW-RE) were consistent with those from IVW-FE, indicating robust results. The funnel plot analysis showed no significant asymmetry, suggesting no publication bias.

**Figure 2.**
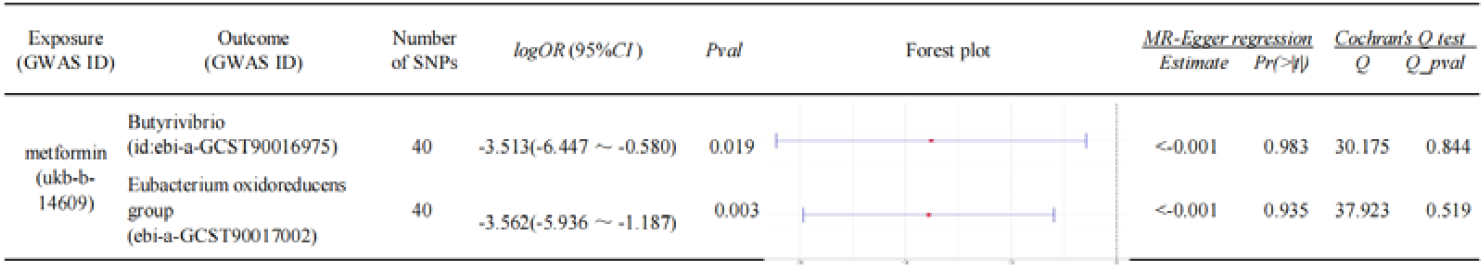
Based on the IVW-FE analysis method, the analysis of the associations between Met and two intestinal microbiota, including *Butyrivibrio* and *Eubacterium oxidoreducens* groups, included the number of SNPs, forest plots of associations, logOR, 95% CI, P-values (pval), intercepts from MR-Egger regression and corresponding horizontal pleiotropy P-values (Pr(>|t|)), and Q-values and corresponding P-values (Q_pval) from Cochran’s Q heterogeneity test.

## 3 Discussion

This study systematically investigated the causal relationships between Met and the abundance of 10 intestinal genera-level microbiota through IVW-FE-based MR analysis, revealing multiple effects and potential mechanisms of Met on the intestinal microbiota.

### 3.1 Bacterial genera showing significant positive causal associations with Metformin

This study found, through MR analysis, that Met showed significant positive causal associations with the abundance of eight intestinal bacterial genera: *Parabacteroides, Bacteroides, Alistipes, Adlercreutzia, Anaerofilum, Oxalobacter, Ruminococcaceae UCG003, and Ruminococcus 1*.

*Parabacteroides, Bacteroides*, and *Alistipes* are important members of the *Bacteroidetes* phylum and key genera in the human intestinal microbiota, playing crucial roles in intestinal health. *Parabacteroides* is enriched in the intestines of centenarians [11] and is closely associated with longevity and intestinal homeostasis. It can regulate the host’s mucosal immune system, enhance intestinal mucosal barrier function [12], and exhibit anti-cancer [13], anti-inflammatory [14], tissue-repairing [15], and host metabolic-regulating effects against obesity [16]. *Bacteroides* can metabolize plant phenolic glycosides to produce aglycones with anti-inflammatory and antibacterial functions, regulating intestinal homeostasis [17], and affect insulin sensitivity and glucose homeostasis through interactions with host metabolic pathways [18]. *Bacteroides* can also produce propionate, which upregulates the expression of the gene encoding leptin, thereby preventing the development of diet-induced obesity [5, 19]. *Alistipes* has a dual role in intestinal health and disease, potentially providing protection against certain diseases [20] while also being a potential pathogenic factor for colorectal cancer [21] and depression [20]. This dual role suggests its complex function in the intestinal microenvironment, possibly influenced by multiple factors.

*Adlercreutzia* has soy isoflavone-metabolizing effects [22] and may exert anti-obesity and anti-diabetic effects through isoflavone metabolism [23]. Additionally, in vitro studies [24] have shown that *Adlercreutzia* exhibits anti-inflammatory properties in human intestinal epithelial cells and hepatocytes by inhibiting the NF-κB pathway. *Oxalobacter* can utilize oxalate as its sole carbon and energy source, reducing intestinal oxalate absorption and lowering the risk of kidney stones [25].

*Ruminococcaceae UCG003* and Ruminococcus 1 play important roles in cellulose degradation and short-chain fatty acid (e.g., acetate, propionate, and butyrate) production. Short-chain fatty acids have various physiological functions, including immunomodulation, reducing inflammatory responses, providing energy, and lowering blood glucose [26]. For example, propionate can inhibit hepatic gluconeogenesis through the GPR43/AMPK signaling pathway, thereby lowering blood glucose levels [27], while butyrate improves blood glucose control by enhancing gut-islet axis signaling (e.g., GLP-1 secretion) [27, 28]. Furthermore, Huang HH et al. [29] observed changes in intestinal microbiota composition after metabolic surgery in diabetic patients and found an increased abundance of *Ruminococcaceae UCG003* post-surgery; Song S et al. [30] indicated through a two-sample MR study that *Ruminococcaceae UCG003* is a protective factor against T2DM.

*Anaerofilum* is an anaerobic bacterial genus [31] with limited research, but based on its anaerobic characteristics and metabolic potential, it is speculated to play roles in organic matter decomposition, energy metabolism, and maintaining ecological balance.

The significant positive causal associations between Met and the above eight intestinal bacterial genera are consistent with some previous studies [32]. Animal experimental studies have shown that Met can increase the abundance of *Bacteroides* and *Parabacteroides* [32-34], and some studies have also indicated that Met increases the abundance of Ruminococcus [35]. However, studies on the correlations between Met and *Alistipes, Adlercreutzia, Anaerofilum, Oxalobacter*, and *Ruminococcaceae UCG003* have not been reported.

Notably, some studies [36, 37] have pointed out that the intestine is another important active target of Met besides the liver. Met can exert its broad physiological effects by influencing the composition of the intestinal microbiota, specifically by regulating intestinal pH, reshaping the structure and function of the intestinal microbiota, to achieve multiple benefits such as improving metabolic status, regulating immune responses, exerting anti-inflammatory and anti-tumor effects, and delaying the aging process [38, 39].

### 3.2 Bacterial genera showing significant negative causal associations with metformin

This study indicated, through two-sample MR analysis, that Met use showed significant negative causal associations with *Butyrivibrio* and *Eubacterium oxidoreducens* groups.

Both *Butyrivibrio* and *Eubacterium oxidoreducens* groups are gram-positive anaerobic bacteria belonging to the Firmicutes phylum. *Butyrivibrio* can produce butyrate through the fermentation of dietary fiber and resistant starch and has strong fiber-degrading capabilities, helping the host better absorb nutrients [40]. Additionally, by producing butyrate, it activates G-protein-coupled receptors (e.g., GPR43 and GPR109a), promoting the secretion of anti-inflammatory cytokines (e.g., IL-10) and reducing intestinal inflammation [41]. It also improves insulin sensitivity, potentially benefiting the prevention and treatment of obesity and type 2 diabetes, and helps alleviate symptoms of inflammatory bowel disease (IBD) by enhancing intestinal barrier function and exerting anti-inflammatory effects. *Eubacterium oxidoreducens* groups can metabolize dietary fiber and, like *Ruminococcaceae UCG003* and Ruminococcus 1, produce short-chain fatty acids [42], which have various physiological functions, including immunomodulation, reducing inflammatory responses, and providing energy [26-28].

The significant negative causal association between Met and *Butyrivibrio* contradicts previous studies (e.g., de la Cuesta-Zuluaga et al. [43]), which indicated that Met increases the abundance of *Butyrivibrio*. This discrepancy may stem from differences in research methods, sample selection, Met dosage, or treatment duration. Studies on the correlation between Met and *Eubacterium oxidoreducens* groups have not been reported. The significant negative causal associations between Met use and *Butyrivibrio* and *Eubacterium oxidoreducens* groups suggest a dual role of Met in regulating the intestinal microbiota.

### 3.3 Limitations and future prospects of the study

#### 3.3.1 Study limitations

Although this study achieved certain results in exploring the causal relationships between Met and intestinal microbiota abundance, some limitations remain.

First, the data for this study were sourced from the IEU OpenGWAS project, which may have biases in terms of ethnicity, geography, and lifestyle, limiting the applicability of the results to other populations. Second, although this study verified the three major assumptions of MR through multiple methods, it is still difficult to fully ensure strict adherence to these assumptions. For example, in the LOO sensitivity analysis, after removing some individual SNPs, the confidence intervals for *Bacteroides, Adlercreutzia, Anaerofilum* id.2053, *Oxalobacter*, and *Ruminococcaceae UCG003* included zero, indicating a significant impact of the removed individual SNPs on the results. Third, this study primarily relied on the IVW-FE method as the main analysis approach, and although other methods were used for auxiliary assessment, different statistical methods have their own assumptions and applicability, potentially introducing bias.

MR analysis can avoid some confounding factors in traditional observational studies but may still be affected by other unconsidered genetic or environmental factors. Potential mediating variables between exposure and outcome may also influence the interpretation of results. Additionally, heterogeneity among different datasets may affect the reliability of the results.

#### 3.3.2 Future research directions

Based on the above study limitations, to gain a deeper understanding of the causal associations between Met and the above 10 intestinal microbiota, future research can proceed in the following two directions:

Conduct large-scale clinical cohort studies to systematically collect host factor data and explore the regulatory effects of Met on the intestinal microbiota under different host conditions through stratified and multifactorial analyses.

Perform intervention experiments in cellular and animal models to further verify the causal effects found in this study and provide more reliable evidence for clinical applications.

## 4 Conclusion

This study, based on MR methods, found significant causal associations between Met and the abundance of multiple intestinal microbiota. Specifically, Met showed positive correlations with the abundance of *Parabacteroides, Bacteroides, Alistipes, Adlercreutzia, Anaerofilum, Oxalobacter, Ruminococcaceae UCG003*, and *Ruminococcus 1*, while showing negative correlations with the abundance of *Butyrivibrio* and *Eubacterium oxidoreducens* groups.

These findings have important biological and clinical implications. First, they further confirm that Met can increase the abundance of *Parabacteroides, Bacteroides*, and

*Ruminococcus*, while presenting conclusions contrary to previous studies regarding the negative correlation between Met and *Butyrivibrio*. Additionally, they provide, for the first time, support for significant positive correlations between Met and *Alistipes, Adlercreutzia, Anaerofilum, Oxalobacter*, and *Ruminococcaceae UCG003*, as well as a significant negative correlation with *Eubacterium oxidoreducens* groups, suggesting a dual role of Met in regulating the intestinal microbiota.

However, this study also has certain limitations. Future research can conduct large-scale clinical cohort studies and cellular/animal model experiments to verify the causal associations found in this study and delve deeper into the specific mechanisms by which Met regulates the intestinal microbiota. It should be noted that the results of this study are based on MR analysis and have certain limitations. Therefore, caution should be exercised in interpreting and applying these results to avoid overinterpretation.

## Data Availability

https://gwas.mrcieu.ac.uk/

